# COMMON ADOLESCENT MENTAL HEALTH DISORDERS SEEN IN FAMILY MEDICINE CLINICS IN GHANA AND NIGERIA

**DOI:** 10.1101/2023.05.04.23289538

**Authors:** JS Kumbet, TIA Oseni, M Mensah-Bonsu, FM Damagum, EA Opare-Lokko, E Namisango, AL Olawumi, OC Ephraim, B Aweh

**Affiliations:** Department of Family Medicine, Geriatric Medicine Sub Division, Federal Medical Centre, Keffi, Nigeria; Department of Family Medicine, Ambrose Alli University, Ekpoma, Nigeria/Lifestyle and Behavioural Medicine Unit, Department of Family Medicine, Irrua Specialist Teaching Hospital, Irrua, Nigeria; Department of Family Medicine, Komfo Anokye Teaching Hospital, Kumasi, Ghana; Department of Family Medicine, Aminu Kano Teaching Hospital, Kano, Nigeria; Department of Family Medicine, Greater Accra Regional Hospital, Accra/Faculty of Family Medicine, Ghana College of Physicians and Surgeons, Accra, Ghana; African Palliative Care Association/College of Health Sciences, Makerere University, Kampala, Uganda; Department of Family Medicine, Alex Ekwueme Federal University Teaching Hospital, Abakaliki, Nigeria; Department of Mental Health and Behavioural Medicine, Irrua Specialist Teaching Hospital, Irrua, Nigeria

**Keywords:** adolescent, mental health, disorder, family physicians, Ghana, Nigeria

## Abstract

**Background:** Mental health disorders among adolescents is on the rise globally. For fear of stigmatization, patients seldom present to mental health physicians. They are mostly picked during consultations with Family Physicians. This study seeks to evaluate the common mental health disorders seen by family Physicians in Family Medicine Clinics in Nigeria and Ghana.

**Methodology:** A descriptive cross-sectional study involving 302 Physicians practicing in Family Medicine Clinics in Nigeria and Ghana who were randomly selected for the study. Data were collected using self-administered semi-structured questionnaire, and were entered into excel spreadsheet before analysing with IBM-SPSS version 22. Descriptive statistics using frequencies and percentages was used to describe variables. Ethical approval was obtained prior to commencement of the study.

**Results:** Of the 302 Physicians recruited for the study, only 233 completed the study, in which 168 (72.1%) practiced in Nigeria and 65 (27.9%) in Ghana. They were mostly in urban communities (77.3%) and tertiary health facilities (65.2%). Over 90% of Family Medicine practitioners attended to adolescents with mental health issues with over 70% of them seeing at least 2 adolescents with mental health issues every year. The burden of mental health disorder was 16% and the common mental health disorders seen were depression (59.2%), Bipolar Affective Disorder (55.8%), Epilepsy (51.9%) and Substance Abuse Disorder (44.2%).

**Conclusion:** Family Physicians in Nigeria and Ghana attend to a good number of adolescents with mental health disorders in their Family Medicine clinics. There is the need for Family Physicians to have specialized training and retraining to be able to recognize and treat adolescent mental health disorders. This will help reduce stigmatization and improve the management of the disease thus reducing the burden.

## INTRODUCTION

The burden of mental health disorders is on the rise globally. In 2019, 1 in every 8 people, or 970 million people around the world were living with a mental disorder, with anxiety and depressive disorders the most common.^1^ In 2020, the amount of people living with anxiety and depressive disorders rose considerably because of the COVID-19 pandemic.^1,2^

Adolescents are group of persons with chronological age 10-19 years of age. Adolescence is a critically important stage of life for mental health and well-being of individuals, not only for the reason that this is when young people acquire autonomy, social interaction, self-control, and rapid learning, but also because the abilities and potentials formed in this period have a direct bearing on their mental health for the rest of their lives.^3^ Although adolescents generally are highly susceptible to mental health challenges, they receive very little attention, especially in developing countries.^4^ Globally, one in seven of adolescents experiences a mental disorder, accounting for 13% of the global burden of disease in this age group with depression, anxiety and behavioural disorders are among the leading causes of illness and disability among them.^5^

According to the World Health Organization, common mental disorders such as depression and anxiety account for the largest proportion of mental, developmental and substance use disorders. Behavioural disorders including attention-deficit and hyperactivity disorder plus conduct disorder are more prevalent among 10-14-year olds, while alcohol and drug use disorders become more common in older adolescence (15-19-year olds). The total number in both age groups combined is 17 million young people, equivalent to almost 20% or one in five of the population in this age group.^3^ A systematic review of studies from Sub-Saharan African countries revealed that 40.8% of adolescents have emotional and behavioral problems, 29.8% have anxiety disorders, 26.9% have depression, 21.5% have PTSD, and 20.8% have suicidal thoughts.^6^

One out of every six young Nigerian aged 15-24 is suffering from poor mental health, according to a report released by the United Nations Children Fund (UNICEF).^7^ A study in Nigeria found that the prevalence of depression among adolescents ranged from 4.2% to 13.6%.^8^ In Ghana, WHO estimated that 650,000 are suffering from a severe mental disorder and a further 2,166,000 are suffering from a moderate to mild mental disorder and the treatment gap is 98% of the total population.^9^ However, the prevalence of mental illness and its burden among adolescents is not known on a national level.^10^

There is a dearth of mental health experts in West Africa. This is worsened by the stigma associated with mental health disorders in the region. The WHO and World Organization of Family Doctors (WONCA) advocates for the integration of mental health services into primary care as the most viable way of closing the treatment gap and ensuring people get the mental health care they need.^11^ The aim of this study was to evaluate the common mental health disorders among adolescents as seen by family Physicians in Family Medicine Clinics in Nigeria and Ghana.

## MATERIALS AND METHODS

### Study Design and Setting

The study was a descriptive cross-sectional study conducted among Family Physicians practising in Nigeria and Ghana from May to September, 2022. It is part of a larger study with some of the findings already published.^12^ The sample size was calculated to be 302 using fisher’s formula and finite correction was made based on the total population of Family Physicians in each country: 1200 and 125 for Nigeria and Ghana respectively. The sample size (302) was proportionately distributed to each country based on their population size at the time of data collection: Nigeria had 254 while Ghana had 48 Family Physicians. The study sites included General Outpatient Clinics of Teaching Hospitals, Specialist, General and District Hospitals and other Primary Healthcare Centers, where Family Physicians practice in both countries. The consenting Physicians were recruited into the study using multi-stage sampling methods. The validated semi-structured self-administered questionnaire was administered to study participants. Other details of the study design and methodology are contained in the published article.^12^ Authors did not have access to information that could identify individual participants during or after data collection as the study was anonymised.

### Statistical analysis

Data were analysed using the Statistical Package for Social Sciences™ (IBM Corp, Armonk, NY, USA) version 22.0. They were presented in tables and were described using frequencies and percentages.

### Ethical Consideration

Ethical approval for the study was obtained from the Health Research Ethics Committee of Irrua Specialist Teaching Hospital, Irrua, Nigeria (ISTH/HREC/20221403/275).

## RESULTS

A total of 233 Family Physicians completed the study (77.2% response rate), in which 65 (27.90%) in Ghana and 168 (72.10%) in Nigeria participated in the study. They worked in facilities that were mainly in the urban setting 180 (77.25%). Majority of the facilities were Tertiary institutions 152 (65.24%), which was either Teaching Hospitals or Federal Medical Centres.

The socio-demographic characteristics are shown in Table 1.

**Table 1:** Socio-demographic characteristics of respondents (N=233)

| VARIABLE | FREQUENCY | PERCENTAGE |
| --- | --- | --- |
| **Country** |  |  |
| Ghana | 65 | 27.90 |
| Nigeria | 168 | 72.10 |
| **Location of Health Facility** |  |  |
| Rural | 53 | 22.75 |
| Urban | 180 | 77.25 |
| **Type of Facility** |  |  |
| General/District Hospital | 55 | 23.61 |
| Teaching Hospital/Federal Medical Centre | 152 | 65.24 |
| Private/Mission Hospital | 26 | 11.15 |

Table 2 shows the adolescent mental health disorders seen in Family Medicine Clinics in Ghana and Nigeria. Over 90% of Family Medicine practitioners attend to adolescents with mental health issues with over 70% of them seeing at least 2 adolescents with mental health issues every year.

**Table 2:** Adolescent mental health disorders seen in primary care clinics (N=233)

| **VARIABLE** | **FREQUENCY** | **PERCENTAGE** |
| --- | --- | --- |
| **Do you attend to adolescents with mental health issues? (n=233)** |  |  |
| Yes | 212 | 90.99 |
| No | 21 | 9.01 |
| **How often do you attend to adolescents with mental health issues? (n=212)** |  |  |
| Often (> 3 patients a year) | 33 | 15.57 |
| Sometimes (2 – 3 patients a year) | 117 | 55.19 |
| Rarely (≤ 1 patient a year) | 62 | 29.24 |

The burden of mental health disorder in this study was 16% and the distribution of the common adolescent mental health disorders seen in Family Medicine Clinics in Ghana and Nigeria are as shown in Table 3. Depression 138 (59.23%) was the most commonly seen disorder followed by Bipolar Disorders 130 (55.79%), Epilepsy 121 (51.93%), and Substance Use Disorders 103 (44.21%) in that order.

**Table 3:** Common Adolescent Mental Health Disorders Seen in Primary Care Clinics (N=233)

| **DISORDER** | **FREQUENCY** | **PERCENTAGE** |
| --- | --- | --- |
| Depression | 138 | 59.23 |
| Substance Use Disorders | 103 | 44.21 |
| Bipolar Disorders | 130 | 55.79 |
| Psychosis | 52 | 22.32 |
| Suicide or Self Harm | 71 | 30.47 |
| Schizoaffective Disorder | 58 | 24.89 |
| ADHD | 39 | 16.74 |
| Epilepsy | 121 | 51.93 |
| Enuresis | 29 | 12.45 |

## DISCUSSION

The study participants and their distribution have already been described in another publication from the study.^12^ Our study showed that 91% of respondents attend to adolescents with mental health issues with over half of them attending to about two to three adolescents with mental health disorders yearly. This is worrisome, it shows that the burden of adolescent mental health in primary care is enormous. If not attended to, it usually leads to major mental problems including suicide and self-harm which have been on the rise among adolescents.^13,14^ It calls for improved capacity in the diagnosis and management of mental health disorders among Family Physicians. This is particularly so as patients hardly present for care as they are generally stigmatized, shunned and denied access to care by their families, caregivers and the society.^15^ The burden of mental health disorders among adolescents in primary care in west Africa in this study (16%) is similar to the Global findings of 14% burden of mental health disorders among adolescents^5^ and 16% burden found by Robert et al in England.^16^ The high burden found in this study can be attributed to the large number of presentation of adolescent to primary care centres as compared to specialist clinics, the fear of stigmatization of mental health disorders and the recent development of subspecialty clinics such as adolescent clinics, and geriatric clinics in General outpatient clinics.^3,10^ Respondents reported a variety of mental health disorders seen. The most common disorder was depression. This was followed by bipolar disorder, epilepsy and substance use disorders with the least common disorders as enuresis, Attention Deficit Hyperactivity Disorder (ADHD), Psychosis and Schizoaffective Disorders. WHO report that 12 billion work hours and 1 trillion US dollars are lost annually to depression and anxiety alone.^17^ On a global scale WHO stated in 2021 that “Depression, anxiety and behavioural disorders are among the leading causes of illness and disability among adolescents”,^18^ which is in tandem with this study having 59.23% of the respondents treating depression. However, a study done in Nigeria and Ghana ^8,19^ had Schizophrenia and Epilepsy as the commonest mental health disorders in Nigeria and Ghana respectively. The above two studies were not done in primary care setting and Providers and stakeholders had limited or no training in adolescent mental health and that could be the reason for the slight difference. The high number of depressions among adolescent can be explained by the high level of poverty and few numbers of specialist to attend to the huge burden of mental health disorder among adolescents.^3^ Suicidal attempt, Suicide and Self Harm may be a complications or an end spectrum of adolescent mental health disorders^13^, in this study 30.47% have suicidal attempt and self-harm which is close to findings of a study done in SSA^6^ which slight difference may be attributable to smaller number of countries involved in our own study.

Bipolar disorder was the second leading mental health disorder identified in this study. This could be due to the fact that the most frequent range of onset of bipolar disorder is between the ages of 14-21 years; which falls with the adolescent and early adult age group.^20^

In this study, epilepsy was placed third among adolescent mental illnesses. This is not surprising given that, compared to all other continents, Africa has the highest prevalence of epilepsy in children and adolescents.^21^ According to a meta-analysis, there are 13–15 new cases of epilepsy for every 1000 people in West Africa.^22^ This was attributed to the high prevalence of parasitic and infectious diseases in Africa.^21^ The influence of epileptogenic recreational drugs and substances; which are commonly used in this age group could also explain the high burden of seizure disorder in them.^21^ The frequent exposure to computer screens, phone screens and flashing lights may also predispose the adolescents to photosensitive epilepsies; which had been reported to be high among them.^23^

Substance use disorder and suicide or self-harm were also prevalent among adolescent in this study. This is similar to the findings of Birhanu *et al* in Ethiopia,^24^ Mavura *et al* in northern Tanzania,^25^ and Volkow *et al* in the US.^26^ The reasons may not be unconnected to the high level of peer influence, risk taking behavior and experimentation with substances due to developmental changes and challenges in adolescence.^24,25^ The high burden of self-harm or suicide in this study could be due to the strong relationship between substance abuse and suicide or self-harm especially, among adolescents and young adults.^27,28^

There is an urgent need for Family Physicians to look out for adolescent mental health issues and address them at the early stage before they progress to more complicated forms. There is also the need for policy makers to increase awareness on the burden of mental health disorders among adolescents and put measures in place to mitigate them.

### Conclusion

The prevalence of mental disorders among adolescents seen by Family Physicians in Family Medicine clinics are high in Nigeria and Ghana. There is the need for Family Physicians to have specialized training and retraining on mental health issues concerning adolescents. There is also a need to have more subspecialty adolescent clinics in Family Medicine Clinics to be able to handle adolescents’ challenges including recognising and treating adolescent mental health disorders easily. There is also the need for high index of suspicion for these disorders in adolescents when they present. Policy makers should also put measures in place to improve awareness and care for patients.

### Limitations

The study was conducted in two countries in West Africa. Though most Family Medicine Clinics in the region are in these two countries, the results still may not be a true representation of the entire region. Also, the study was conducted among doctors. However, relatively most primary health care centres in the region are run by primary care nurses, community health officers and community health extension workers. These categories of primary care providers were not included in the study even though they attend to most of the patients presenting to primary care facilities in these regions.

## Data Availability

All relevant data are within the manuscript and its Supporting Information files.

## Author Contribution

SK, OTIA, MBM, DF, OLE and EO conceived the idea, designed the study and conducted the study. NE analysed the data. All authors wrote the manuscript, edited and revised the manuscript as well as approved the final copy of the manuscript.

## Acknowledgement

We thank Afriwon Research Group ARG of Afriwon Renaissance for bringing young researchers in Africa together and providing the platform from which the research was conducted.

